# Device-measured physical activity and sedentary behavior during office vs. remote workdays among hybrid workers

**DOI:** 10.1101/2025.11.11.25339977

**Authors:** Tuija Leskinen, Miika Tuominen, Kristin Suorsa, Jesse Pasanen, Sanna Pasanen, Vilma Iisalo, Virpi Ruohomäki, Katja Pahkala, Olli J Heinonen, Sari Stenholm

**Affiliations:** Department of Public Health, University of Turku and Turku University Hospital, Finland; Centre for Population Health Research, University of Turku and Turku University Hospital, Finland; Finnish Institute of Occupational Health, Helsinki, Finland; Research Centre of Applied and Preventive Cardiovascular Medicine, University of Turku, Finland; Paavo Nurmi Centre and Unit for Health and Physical Activity, University of Turku, Finland; Research Services, Turku University Hospital and University of Turku, Finland

**Author notes:** Correspondence: Dr. Tuija Leskinen, Department of Public Health, FI-20014 University of Turku, Finland.

**Keywords:** workday, physical activity, sedentary behavior, remote work, hybrid work

## Abstract

**Objectives:** Daily sedentary time has been found to be higher during remote workdays compared to office workdays, but current evidence relays on studies from pandemic times. This study aimed to compare device-measured physical activity and sedentary behavior between office and remote workdays, worktimes, and non-worktimes among Finnish hybrid workers.

**Methods:** Overall, 97 university employees (84% women, mean age 41.6 (SD 10.5) years) wore Fibion SENS sensor on their thigh for 7 consecutive days and provided at least one remote and one office workday. Sedentary time (SED), light physical activity (LPA), and moderate-to-vigorous physical activity (MVPA) were measured for remote and office workdays, worktimes, and non-worktimes. A linear mixed model was used to study the within-individual differences.

**Results:** Remote workdays accumulated 44 min (95% confidence interval (CI) 25-64) more SED, 29 min (95% CI 10-48) less LPA, and 15 min (95% CI 11-20) less MVPA compared to office workdays. Remote worktime included 31 min (95% CI 15-48) more SED, 24 min (95% CI 8-40) less LPA, and 7 min (95% CI 5-10) less MVPA compared to the office worktime. Non-working hours accumulated more SED and less MVPA during remote vs. office workday.

**Conclusions:** Remote workdays included less physical activity and more sedentary behavior compared to office workdays, especially during working hours. Because of the popularity of remote work, strategies to promote physical activity and reduce sedentary behavior during remote work are warranted.

**What is already known on this topic:** Remote work is a popular way of working, but it may increase daily sedentary time.

**What this study adds:** Higher levels of sedentary behavior and lower physical activity during remote vs. office workdays derived mainly from morning and working hours.

**How this study might affect research, practice or policy:** Actions and interventions to increase physical activity and reduce sedentary behavior during remote working hours are needed.

## Introduction

Daily physical activity is accumulated during worktime, leisure time, and household chores. It is well-known that occupation contributes to daily total physical activity[1]. Unlike manual workers, who accumulate most of their daily physical activity during working hours, office employees spend the majority of their worktime sedentary[2]. Sedentary behavior, i.e., wake time sitting and laying, is a well-known risk factor for metabolic disturbances[3,4]. Therefore, evidence-based interventions and actions, e.g., the use of sit-stand desks, have been applied to reduce sitting time in office environments[5–7].

After the COVID-19 pandemic, remote work has remained a common way of working[8]. According to recent statistics, over one fifth of the US citizens[9] and Europeans[10] work at least partly remotely, with Finland having similar percentages for remote work[11]. Previous survey-based studies have shown that working from home during the pandemic was associated with higher daily sedentary time than working at the office[12], thus raising a health concern of increased sedentary time[13]. The higher sedentary time while working from home has been explained by the lack of active commuting and social interaction, and by limited space in home offices[12]. Because of these differences in work environments and daily behavior, the current office-based actions and tools to increase physical activity or decrease sedentary time may not be suitable for remote work[14]. Therefore, more studies on physical activity behavior during remote workdays are needed[13].

Modern device-based measurements have enabled the collection of accurate data on physical activity and sedentary behavior during entire 24-hour day, including working hours[15,16]. To date, only a few accelerometer-based studies on physical activity behavior among hybrid workers have been published. These studies have shown that remote workdays accumulate more in-bed time[17], higher daily sedentary time[18,19], and less daily moderate to vigorous physical activity[20] when compared to office workdays. However, these previous studies have been conducted with rather small sample sizes and during the pandemic restrictions, which overall reduced daily physical activity[21]. Also, the existing findings are mostly reliant on daily level assessments, which does not allow for distinguishing between physical activity and sedentary behavior accumulated during working hours vs. non-working hours. To fill this gap, we examined the within-individual differences in daily total, worktime, and non-worktime physical activity and sedentary behavior between office and remote workdays among Finnish university employees. Participants had high degree of flexibility to decide whether to work remotely or at their employer’s premises. Therefore, they are called here as hybrid workers.

## Methods

The Daily Physical Activity among Hybrid Workers (WORKDAY study) aimed to compare physical activity and sedentary behavior between office and remote workdays. Study participants were recruited from the campus area of the University of Turku, University hospital, and University of Applied Sciences, Finland, by advertising the study in the internal bulletins, units’ email lists, and campus buildings’ noticeboards during May 2023 to June 2024. Eligible participants were those office employees who were able to work both at the office and remotely, i.e., hybrid work. A link/QR code for the registration questionnaire was offered in the advertisement. The registration questionnaire included a description of the study with a choice of participation (yes/no). If agreed, the respondent was forwarded to the participant information sheet. After filling in their contact information, the respondent was asked to select a preferred measurement week.

Overall, 108 persons answered the registration questionnaire, of which four responses were direct refusals. Of those agreeing, 99 participants (84% women) were available for the measurements and responded to the study questionnaire. Five participants dropped out due to scheduling issues. The analytical sample of 97 hybrid workers consisted of participants who provided at least one valid remote workday and one valid office workday.

The WORKDAY study has a positive ethical review statement from the Ethics Committee for human sciences at the University of Turku (13/2023). All participants gave their written informed consent before the measurements. A power analysis to determine the minimum sample size of N=88 was calculated for a 30-minute (SD 99[19]) difference in daily sedentary time, with a significance level of 0.05 and desired power of 0.80.

### Measurement of physical activity and sedentary behavior

Physical activity and sedentary behavior were measured with the Fibion SENS Motion System, which includes a triaxial SENS motion® sensor, a smartphone application, and a web-based application[22]. The Fibion SENS sensor was initialized to record acceleration with a 12.5 Hz sampling frequency. Research personnel met each participant in person and provided both verbal and written instructions on how to wear the sensor. The participants were instructed to fasten the sensor with an adhesive waterproof film dressing directly to the skin on the midway of the thigh, approximately 5−10 cm above the knee joint. The participants were asked to wear the sensor continuously for 24 hours per day for at least seven consecutive days, including during water-based activities, but to remove it for sauna or in case of skin irritation. The measurement period was requested, but not restricted, to include two office workdays and two remote workdays. The participants were also instructed to fill in a daily log during their measurement period to record daily work modes (office, remote, combined), daily working hours, and in-bed and out-bed times. After the measurement period, the participants returned the sensors and the daily logs to pointed return boxes located within the University buildings or via mail. The sensor did not provide any feedback to the participants while measuring; however, after data processing, each participant received detailed feedback on their daily sedentary behavior and a suggestion for actions.

The raw data from the sensors were downloaded via Bluetooth using a smartphone application and transmitted to a secure web server for storage and later analysis. Data were sampled in 5-s epochs and automatically categorized into time spent in the predefined activity categories using a validated built-in algorithm[22]. The activity categories included sitting or laying awake as sedentary behavior (SED); standing, sporadic walking, and continuous light walking as light physical activity (LPA); biking, moderate intensity activity (e.g., brisk walking), and high intensity activity (e.g., running) as moderate to vigorous physical activity (MVPA). Total steps taken were calculated for each epoch.

A valid measurement day was defined as a minimum of 10 hours of sensor wear time during waking hours. Sleep time was estimated using in-bed and out-of-bed times from daily logs. The non-wear time of the sensor was defined when activity categorization was not possible and excluded using the predefined category called “no data”[22]. The mean time spent in different activity categories and the number of steps were calculated for valid office and remote workdays, and separately for working hours and non-working hours (i.e., time outside the reported working hours) within each workday type. Valid workdays including both office and remote worktimes were not included in the analysis (n=19 days, 4% of all valid workdays). The hourly total physical activity minutes (LPA+MVPA) was used to illustrate the average accumulation of physical activity during typical waking hours (from 6 to 23 hours) for both office and remote workdays.

### Participant characteristics

Age, sex, occupational title, self-rated health, self-reported height and weight, living alone (yes/no), and the number of minor children were acquired with a web-based study questionnaire. Occupational titles were coded according to the ISCO classification[23] into: professionals (ISCO 1-2) and associate professionals and service workers (ISCO 3-5). Self-rated health was assessed by a 5-point scale: good, rather good, average, rather poor, poor. Body Mass Index (BMI) was calculated from self-reported body weight and height as kg/m^2^ and classified according to WHO into five categories: underweight, normal, overweight, moderate obese, severe obese[24].

### Statistical analysis

The characteristics of the participants are presented as mean values and standard deviations (SD) for the continuous variables and as frequencies and percentages for the categorical variables. The within-individual differences in mean levels of SED, LPA, MVPA, and steps between office and remote workdays, worktime, and non-worktime were examined using linear mixed models, adjusted for sensor wear time. The results are given as mean estimates and mean difference estimates with their 95% confidence intervals (CI). The SAS Software 9.4 was used for the statistical analyses (SAS Institute Inc., Cary, NC).

## Results

The mean age of the study participants was 41.6 (SD 10.5) years, ranging from 25 to 65 years (Table 1). Overall, 84% were women, 85% had a professional occupation, and 55% had a normal BMI. Of the participants, 63% had good self-reported health, 25% were living alone, and 60% did not have minor children. Measurement data included mean of 2.4 (SD 0.8) valid office workdays and 2.6 (SD 0.8) valid remote workdays. The majority (89%) of the remote workdays took place at home. The observed levels of SED, LPA, and MVPA for office and remote workdays are shown in Supplementary Figure 1.

**Table 1.** Characteristics of the analytical sample of hybrid workers (n=97).

|  |  |
| --- | --- |
|  | Mean (SD) |
| Age, years | 41.6 (SD 10.5) |
|  | N (%) |
| Women | 81 (84%) |
| Occupation |  |
| Professionals | 82 (85%) |
| Associate professionals<br>and service workers | 15 (15%) |
| BMI classes |  |
| Underweight | 0 (0%) |
| Normal | 53 (55%) |
| Overweight | 31 (32%) |
| Moderate obese | 10 (10%) |
| Severe obese | 3 (3%) |
| Self-reported health |  |
| Good | 60 (63%) |
| Rather good | 27 (28%) |
| Average | 9 (9%) |
| Rather poor | 0 (0%) |
| Poor | 0 (0%) |
| Living alone | 24 (25%) |
| No minor children | 58 (60%) |

SD=standard deviation

Table 2 presents comparisons for daily total, worktime, and non-worktime physical activity and sedentary behavior between office and remote workdays. On average, participants accumulated SED 9 hours 42 minutes on office workdays compared to 10 h 27 min on remote workdays (mean difference 44 min, 95% CI 25 to 64). In addition, remote workdays included less LPA (mean diff. 29 min, 95% CI 10 to 48), less MVPA (mean diff. 15 min, 95% CI 11 to 20), and fewer steps (mean diff. 1067, 95% CI 256 to 1878) compared to office workdays. Figure 1 illustrates the hourly accumulation of total physical activity (LPA+MVPA) during office and remote workdays. Although these two patterns were rather similar in shape, more physical activity was accumulated during office workdays’ morning hours.

**Table 2.** Within-individual comparisons for daily total, worktime, and non-worktime behaviors between office and remote workdays.

|  | **Office workdays** | **Remote workdays** | **Difference between office and remote workdays** |
| --- | --- | --- | --- |
|  | Mean min (95% CI) | Mean min (95% CI) | Mean min (95% CI) |
| **Wear time** |  |  |  |
| Daily total | 972 (963 to 982) | 961 (951 to 970) | -12 (-23 to -0.6) |
| Worktime | 458 (447 to 470) | 459 (447 to 470) | 0.4 (-15 to 15) |
| Non-worktime | 495 (478 to 512) | 493 (476 to 510) | -2 (-25 to 21) |
| **Sedentary behavior** |  |  |  |
| Daily total | 582 (559 to 606) | 627 (603 to 650) | 44 (25 to 64) |
| Worktime | 300 (283 to 316) | 331 (315 to 347) | 31 (15 to 48) |
| Non-worktime | 275 (262 to 288) | 288 (275 to 301) | 14 (3 to 25) |
| **Light physical activity** |  |  |  |
| Daily total | 337 (314 to 360) | 308 (285 to 331) | -29 (-48 to -10) |
| Worktime | 146 (129 to 162) | 122 (105 to 138) | -24 (-40 to -8) |
| Non-worktime | 186 (174 to 198) | 180 (168 to 191) | -6 (-16 to 4) |
| **Moderate-to-vigorous physical activity** |  |  |  |
| Daily total | 47 (43 to 52) | 32 (27 to 36) | -15 (-20 to -11) |
| Worktime | 13 (11 to 15) | 6 (4 to 7) | -7 (-10 to -5) |
| Non-worktime | 33 (29 to 37) | 26 (22 to 30) | -7 (-12 to -3) |
| **Number of daily steps** |  |  |  |
| Daily total | 14799 (13930 to 15667) | 13732 (12869 to 14595) | -1067 (-1878 to -256) |
| Worktime | 4722 (4299 to 5145) | 3980 (3563 to 4398) | -742 (-1197 to -286) |
| Non-worktime | 9853 (9163 to 10543) | 9474 (8789 to 10158) | -380 (-1062 to 303) |

CI=confidence interval
Models were adjusted for daily, worktime, and non-worktime sensor wear time according to the comparison.

**Figure 1.**
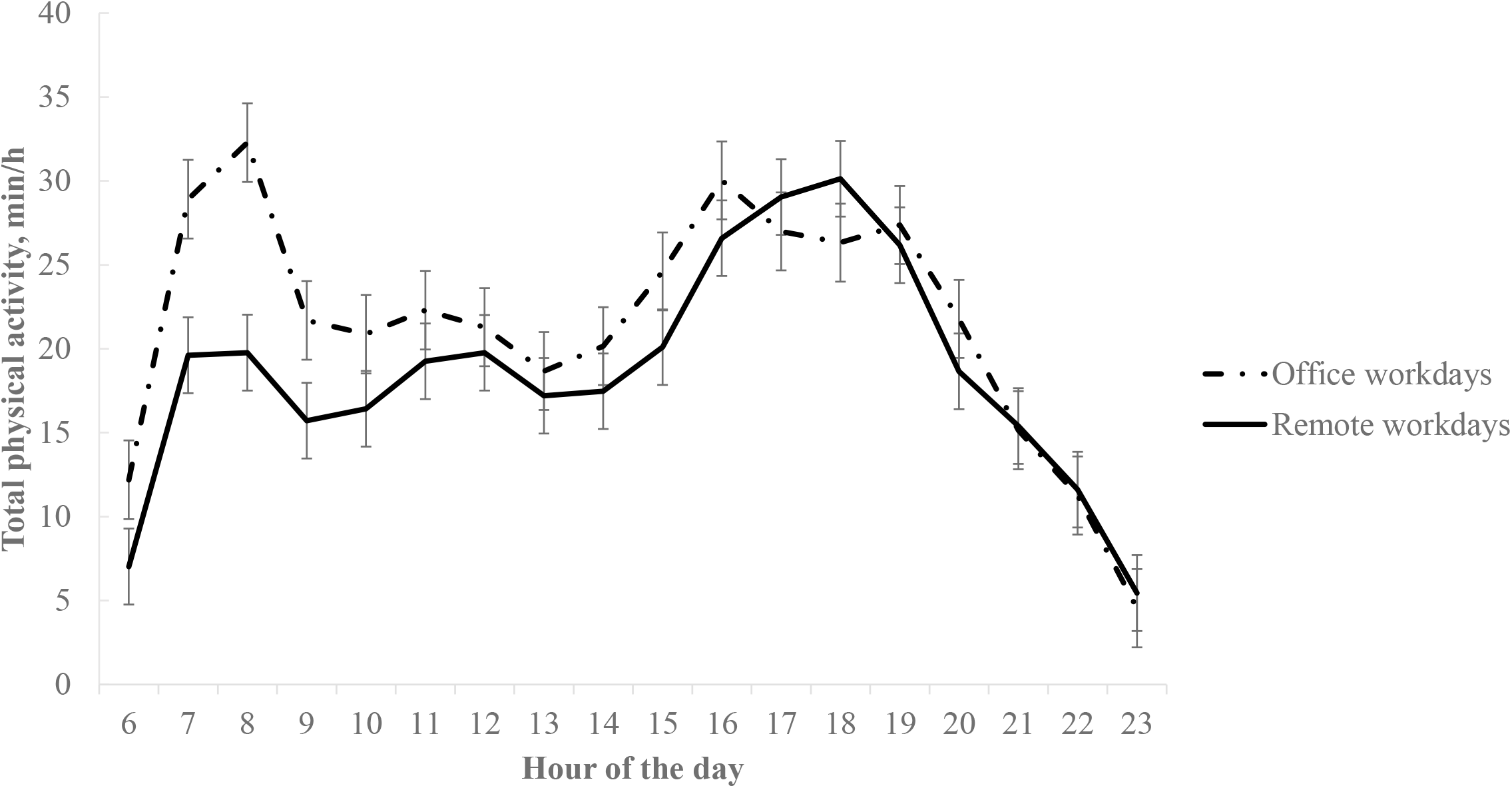
Hourly accumulation of total physical activity time (minutes) during office and remote workdays among hybrid workers.

When focusing on the differences within the working hours, the office worktime included 5 h 0 min of SED in comparison to 5 h 31 min during remote workdays (mean diff. 31 min, 95% CI 15 to 48). Remote worktime included also less LPA (mean diff. 24 min, 95% CI 8 to 40), less MVPA (mean diff. 7 min, 95% CI 5 to 10), and fewer steps (mean diff. 742, 95% CI 286 to 1197) compared to the office worktime. No differences in worktimes between office and remote workdays were observed (Table 2).

During non-working hours, remote workdays included 14 minutes (95% CI 3 to 25) higher SED and lower MVPA (mean diff. 7 min, 95% CI 3 to 12) compared to non-working hours in office workdays, whereas the amount of non-worktime LPA and steps did not differ between the workday types (Table 2).

## Discussion

This study aimed to compare daily total, worktime, and non-worktime physical activity and sedentary behavior between office and remote workdays among Finnish hybrid workers. Our results showed that remote workdays accumulated 44 minutes more SED compared to office workdays. The difference was especially seen during working hours, as 31 minutes more SED was accumulated during remote vs. office worktime. Similarly, remote workdays accumulated 29 minutes less LPA and 15 minutes less MVPA compared to office workdays. The difference in physical activity was seen during morning hours and within the reported working hours.

Our finding of higher SED during remote vs. office workdays is in line with previous, pandemic times accelerometer-based studies among hybrid workers[19,20]. The 31-minute difference in worktime SED is similar to wrist-worn accelerometer studies showing 20 to 26 minutes more SED when working remotely vs. office among hybrid workers[19,25]. Similarly, the lower LPA and MVPA levels during remote vs. office workdays complement previous accelerometer-based findings among hybrid workers [19,20]. The added contribution of the current study was that we were able to examine working and non-working time separately and observed that the difference in LPA and steps accumulated during worktime, whereas the MVPA difference was seen during both worktime and non-worktime. The lower non-worktime MVPA during remote vs. office workdays may refer to the lack of physical activity related to commuting. Unfortunately, we were not able to differentiate commuting activity in our study. However, hourly patterns showed that the difference in physical activity between office and remote workdays was seen during the morning hours when commuting to work usually occurs. This observation suggests that the physical activity that is lost due to lack of commuting is not likely to be replaced during the day when working remotely.

There are several possible explanations for the lower physical activity and higher SED during remote working hours. The home environment, in which most of the remote work is done, may demand shorter transitions to e.g., lunch, coffee, or printer compared to office environments[12], in which these transitions usually accumulate more physical activity[26]. The absence of sit-stand desks at home may, in turn, make it more challenging to work while standing, and limit opportunities for taking breaks from sitting, particularly during tasks requiring on-camera attendance, such as online meetings[14]. Moreover, in desk-based jobs, sitting is likely to occur as an automatic by-product of carrying out work tasks[27]. As the working hours at home are likely to be less structured and include fewer external interruptions, the sitting automaticity may be reinforced through increased immersion in work tasks[14,28]. Thus, prolonged sitting bouts might become longer and more frequent while working from home, particularly during concentrated work.

Breaking up prolonged sitting has been shown to offer benefits to e.g., postprandial glucose[29–31] and mental wellbeing, body composition, and blood pressure[3], particularly when breaks from sitting are spent physically active. Decreasing worktime sedentary behavior may also improve work performance, mood, and work satisfaction, and reduce musculoskeletal pain[32]. Previous interventions to break up worktime sitting, e.g., with sit-stand desks or digital prompts, have been shown to be effective in reducing SED in office environments[6,33]. However, there is a lack of evidence on whether these office-based tools are applicable, feasible, or effective in reducing sitting and/or increasing physical activity at home or other remote work environments[13,14,33]. In addition, education, social support and modeling, e.g., breaking up sitting during meetings by the managers, are suggested to play an important role in reducing SED in office[6] but also when working from home[14]. In fact, support from co-workers has been shown to lead successful interventions in office-based studies[7], whereas support from employers or managers may be more crucial when working remotely[34].

Taken together, there are multiple aspects that need to be taken into consideration in future recommendations and interventions aiming to increase physical activity and decrease sedentary behavior among remote workers. Successfully reducing worktime sitting is likely to require multi-component interventions, with focus on behavioral self-regulation, environmental opportunities, and social norms and practices associated with break-taking[28,34]. It is also noteworthy that promoting MVPA has proven difficult at office-based interventions[5], suggesting that overall new strategies to increase MVPA among office employees are needed. Working from home might hold potential for promoting MVPA, given that common barriers to increase MVPA at the office, such as workplace culture and lack of flexibility[35], may not be as relevant at home. At best, more flexible working hours, absence of social barriers, and e.g., easier access to outdoors when working from home could provide opportunities for longer and higher-intensity active breaks or to increase time in recreational or household activities[36]. This could further serve to replace the physical activity that appears to be lost due to lack of commuting.

Major strength of our study is that it was conducted among current hybrid workers, enabling within-individual comparison of physical activity behavior between office and remote workdays after pandemic times. With daily logs, we were able to identify daily working hours to study worktime physical activity and SED apart from non-working hours. As a methodological strength, we used 24/7 thigh-worn sensors to sensitively capture postural changes required for the separation of different activities[22]. The generalizability of our results may be limited as we recruited mostly university employees, among whom the level of education and work-related autonomy might be relatively high[37]. However, there is also role-specific differences in sitting time as well as in possibilities to break up sitting among employees in the academic environment[38].

## Conclusions

Remote workdays accumulate lower physical activity and higher sedentary time than office workdays among hybrid workers. The differences are mostly accrued during morning hours and within worktime. Due to the popularity of remote work, these findings call for actions to increase physical activity and reduce sedentary behavior during remote workdays.

## Acknowledgements

The WORKDAY study warmly thanks the research personnel Kati Pyykkö and Roosa Latvaniemi and all the study participants.

## Contributorship statement

Tuija Leskinen, Sari Stenholm, and Miika Tuominen designed the study and data collection. Katja Pahkala and Olli J Heinonen contributed to accelerometry data acquisition. Kristin Suorsa, Jesse Pasanen, Sanna Pasanen, Vilma Iisalo, and Virpi Ruohomäki participated to data curation. Tuija Leskinen conducted the analyses with the help of Kristin Suorsa and Jesse Pasanen. Tuija Leskinen drafted the manuscript and is the guarantor. All authors contributed to data interpretation, revised article critically, and approved the final version of the manuscript.

## Ethics statement

The WORKDAY study has a positive ethical review statement from the Ethics Committee for human sciences at the University of Turku (13/2023).

## Data availability

Anonymized partial datasets are available upon reasonable request.

---

**Supplementary Figure 1.**
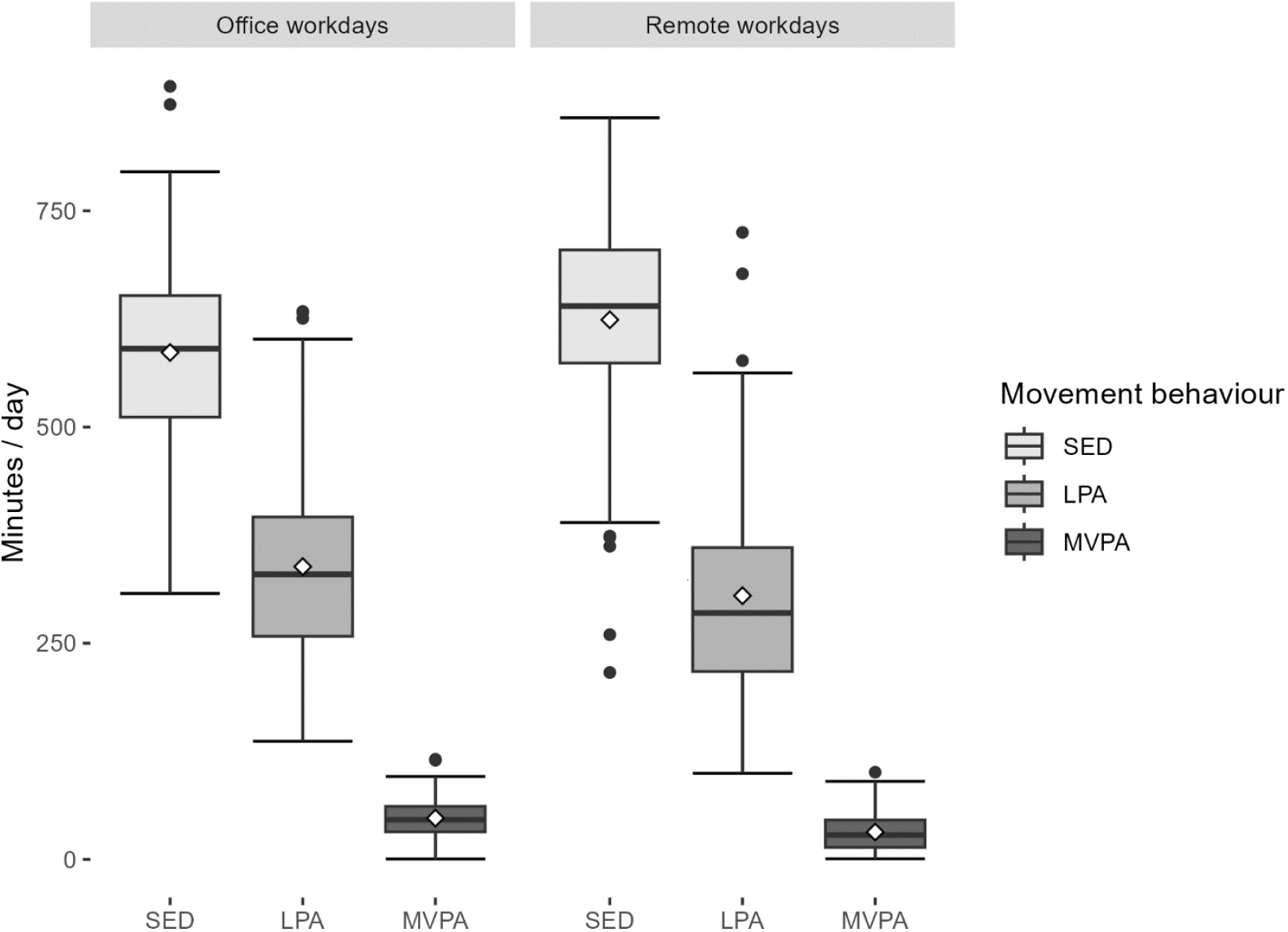
Boxplots of the observed daily minutes of sedentary time (SED), light physical activity (LPA), and moderate to vigorous physical activity (MVPA) for office and remote workdays among the hybrid workers.

